# Study Data Element Mapping: Feasibility of Defining Common Data Elements Across COVID-19 Studies

**DOI:** 10.1101/2020.05.19.20106641

**Authors:** P Mathewson, B Gordon, K Snowley, C Fennessy, A Denniston, NJ Sebire

**Author notes:** **Correspondence:**, Prof NJ Sebire, HDRUK, Gibbs Building, 215 Euston Road, London NW1 2BE, UK.

## Abstract

**Background:** Numerous clinical studies are now underway investigating aspects of COVID-19. The aim of this study was to identify a selection of national and/or multicentre clinical COVID-19 studies in the United Kingdom to examine the feasibility and outcomes of documenting the most frequent data elements common across studies to rapidly inform future study design and demonstrate proof-of-concept for further subject-specific study data element mapping to improve research data management.

**Methods:** 25 COVID-19 studies were included. For each, information regarding the specific data elements being collected was recorded. Data elements collated were arbitrarily divided into categories for ease of visualisation. Elements which were most frequently and consistently recorded across studies are presented in relation to their relative commonality.

**Results:** Across the 25 studies, 261 data elements were recorded in total. The most frequently recorded 100 data elements were identified across all studies and are presented with relative frequencies. Categories with the largest numbers of common elements included demographics, admission criteria, medical history and investigations. Mortality and need for specific respiratory support were the most common outcome measures, but with specific studies including a range of other outcome measures.

**Conclusion:** The findings of this study have demonstrated that it is feasible to collate specific data elements recorded across a range of studies investigating a specific clinical condition in order to identify those elements which are most common among studies. These data may be of value for those establishing new studies and to allow researchers to rapidly identify studies collecting data of potential use hence minimising duplication and increasing data re-use and interoperability

## Introduction

Since the development of the global COVID-19 emergency in early 2020, more than 2000 clinical studies have now been registered investigating various aspects of the disease.^1^ Ideally, all of these studies would learn from each other, use common data standards, definitions and formats and allow maximum interoperability and data pooling. In addition, for new studies being instituted to address specific aspects of COVID-19 or specific interventions, the data elements collected would ‘reuse’ existing data element components from existing studies, only adding specific additional variables where required. In practice, in part due to the rapidity of their inception and development, and in part due to longstanding cultural habits, it is likely that most studies continue to be established ‘de-novo’ with little focus on data reuse and interoperability, these being considered secondarily once data collection is complete.

The aim of this study was to identify a small selection of major national and/or multicentre clinical COVID-19 studies in the United Kingdom to examine the feasibility and outcomes of documenting specific data elements being collected, in order to identify the most frequent data elements common across studies to provide an evidence-based list of relatively ranked elements which may inform future study design. In addition, these findings could act as a proof-of-concept for further subject-specific study data element mapping more widely across the healthcare landscape as a contributing factor to improve research data design and management and support FAIR data.^2,3^

## Methods

We selected 25 major studies for inclusion, representing a cross-section of large multicentre and/or National studies set up in the first weeks of the COVID-19 pandemic in the United Kingdom.^4–7,8,9,10,11–13,14,15,16,17,18,19,20,21,22–24,25,26^ There was no attempt to identify specific COVID-19 disease areas but studies were selected to include the major studies documented on the UK National Institute for Health Research (NIHR) and Health Research Authority (HRA) websites during April 2020.

For each study, either the specific study website, (if available), or study information available from other sources such as trial registers, were accessed in order to identify the specific data elements being collected in the study. This information was either available from dedicated study data element data documentation in association with data dictionaries and coding schemes used, directly from the study specific eCRF (electronic case report forms), or from study protocols where neither of the previous sources were available. We arbitrarily started by mapping data elements from the UK CHESS study,^4^ recording the data elements, definitions if available and any specific comments. Following this, data elements from additional studies were manually recorded, either aligning to the pre-existing list where appropriate, or adding new elements where required. Data elements collated were arbitrarily divided into categories, such as demographics, investigations, medical history for ease of visualisation and discussion. For some elements subjective judgement was required, for example some studies may have included an element as an inclusion or exclusion criteria but not explicitly documented this in the eCRF.

Following the recording and collation of elements across all studies, the elements from categories which were most frequently and consistently recorded across studies were presented in list form and using sunburst and treemap charts in order to visually present the major data elements in relation to their relative commonality. Analysis was performed using Microsoft Excel (Microsoft Corp, Seattle). Research ethics committee approval was not required for this work.

## Results

Across the 25 studies, 261 data elements were recorded in total. The most frequently recorded 100 data elements were identified across all studies and are provided in Table 1 and Figures 1 and 2, with relative frequencies in terms of proportion of studies collecting such data element. The groups with the largest numbers of common elements included demographics, admission criteria, medical history and investigations. Mortality and need for specific respiratory support were the most common outcome measures, but with specific studies including a range of other outcome measures such as length of stay, oxygen requirements etc. Given that many studies were investigating specific interventions or treatments, these of course would be highly relevant to the specific study but unlikely to be common across many studies, hence are not listed here.

**Table 1.** Most frequent 100 COVID-19 study data elements presented as list form in relation to major category

| **Element type** | **Entity** |
| --- | --- |
| Demographics | Age |
| Demographics | Sex |
| Demographics | Pregnant |
| Demographics | Healthcare worker |
| Demographics | NHS number |
| Demographics | Hospital number |
| Demographics | Ethnicity |
| Demographics | Weight/height |
| Demographics | Travel to infected area |
| Demographics | Date of birth |
| Demographics | Post code stem |
| Demographics | Ethnicity |
| Demographics | Contact with known infected case |
| Demographics | Organisation / HOSP NAME |
| Demographics | GP practice |
| Admission | Mortality |
| Admission | Respiratory support |
| Admission | Date of admission |
| Admission | Admitted to ICU |
| Admission | Date of admission to ICU |
| Admission | Admission COVID-19 related |
| Admission | Length of hospital stay |
| Admission | Outcome date |
| Admission | Duration of mechanical ventilation |
| Admission | Date of symptoms onset |
| Admission | Length of ICU stay |
| Admission | Duration of oxygen suppl |
| Admission | Symptom duration (days) |
| Admission | Admitted from |
| Admission | Major admission code |
| Admission | mortality in ICU |
| Admission | mortality in hospital stay |
| Medical History | Cardiac disease / CHD |
| Medical History | Chronic pulm disease (not asthma) |
| Medical History | Asthma |
| Medical History | Chronic kidney disease |
| Medical History | Diabetes |
| Medical History | Hypertension |
| Medical History | Chronic neurological disorder |
| Medical History | Chronic haematological disorder |
| Medical History | AIDS/HIV |
| Medical History | Smoking |
| Medical History | Mod/sev liver disease |
| Medical History | Malignant neoplasm |
| Medical History | Rheumatological disease |
| Medical History | Dementia |
| Medical History | Medical History NOS |
| Medical History | Medication history / Drugs history |
| Medical History | immunosuppressed |
| Medical History | Obesity |
| Medical History | Malnutrition |
| Investigations | SARS-COV-2 test |
| Investigations | CXR / imaging |
| Investigations | COVID Lab test date |
| Investigations | Creatinine |
| Investigations | Lymphocyte count |
| Investigations | WBC count |
| Investigations | CRP level |
| Investigations | Liver function tests |
| Investigations | Specimen type |
| Investigations | Lab test method |
| Investigations | Blood gas parameters |
| Investigations | Haemoglobin concentration |
| Investigations | Platelet count |
| Investigations | Other microbiology |
| Investigations | Sodium |
| Investigations | Potassium |
| Investigations | RSV |
| Investigations | Adenovirus |
| Symptoms | Cough |
| Symptoms | Fever |
| Symptoms | Sore throat |
| Symptoms | Myalgia |
| Symptoms | Diarrhoea |
| Symptoms | decreased exercise tolerance |
| Symptoms | Cough with sputum |
| Symptoms | Headache |
| Symptoms | Vomiting/nausea |
| Symptoms | Fatigue |
| Symptoms | Abdominal pain |
| Clinical features | Respiratory rate |
| Clinical features | Temp |
| Clinical features | Dyspnoea |
| Clinical features | Pneumonia (clinical) |
| Observations | O2 sat / oxygen saturations |
| Observations | pAO2 |
| Observations | Heart rate |
| Observations | Blood pressure |
| Observations | Clinical symptom score eg<br>APACHEII |
| Observations | Mild/Mod/severe severity |
| Observations | Glasgow coma score |
| Treatment | Antiviral therapy |
| Treatment | Inotropic support |
| Treatment | Other prescribing |
| Treatment | Therapy at discharge |
| Treatment | Dialysis |
| Treatment | Intubation |
| Treatment | Tracheostomy |
| Treatment | Antibiotics |
| Treatment | FiO2 |

**Figure 1.**
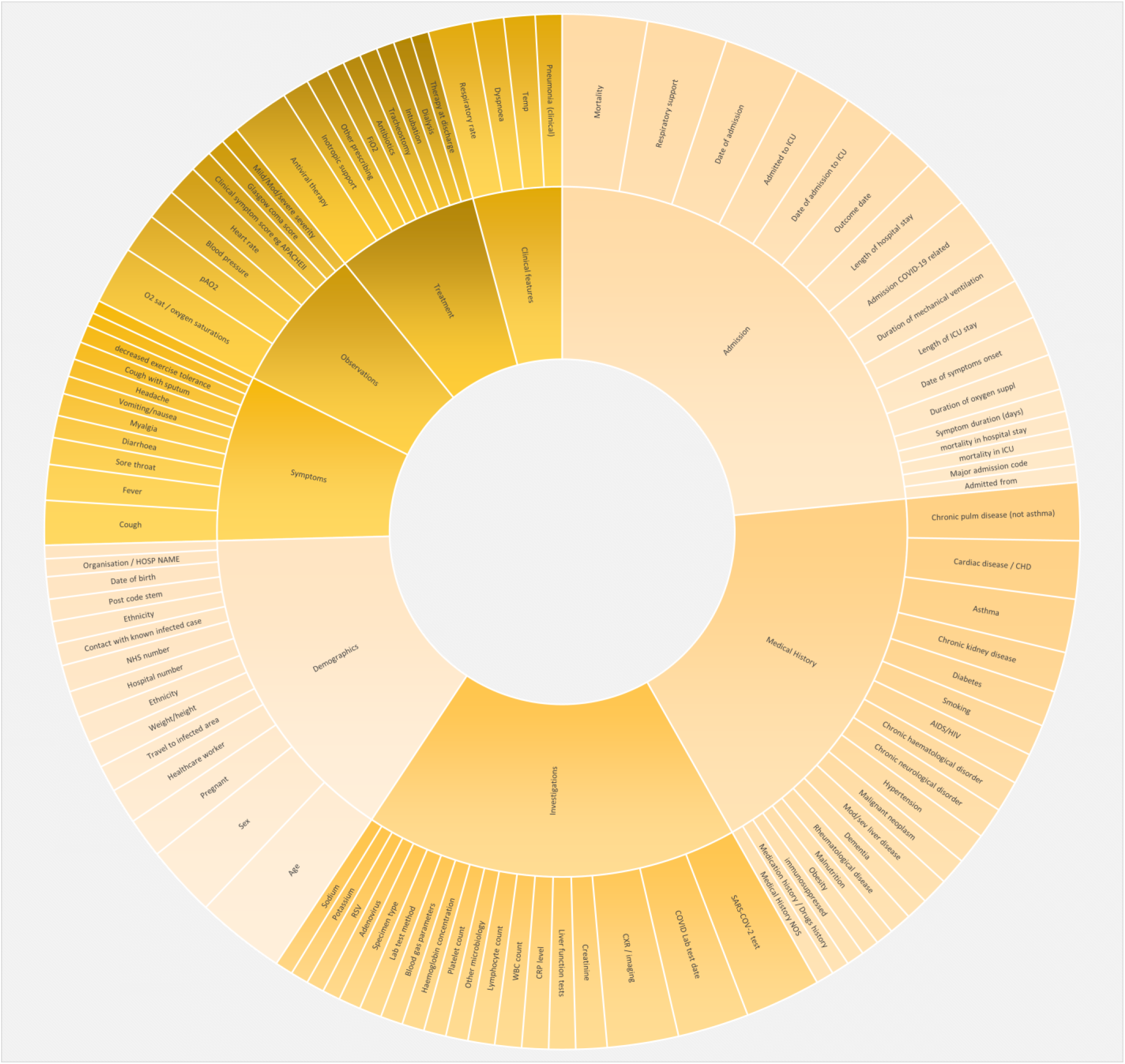
Most frequent 100 COVID-19 study data elements presented as Sunburst chart in which the relative size of the segments represents relative frequency across studies.

**Figure 2.**
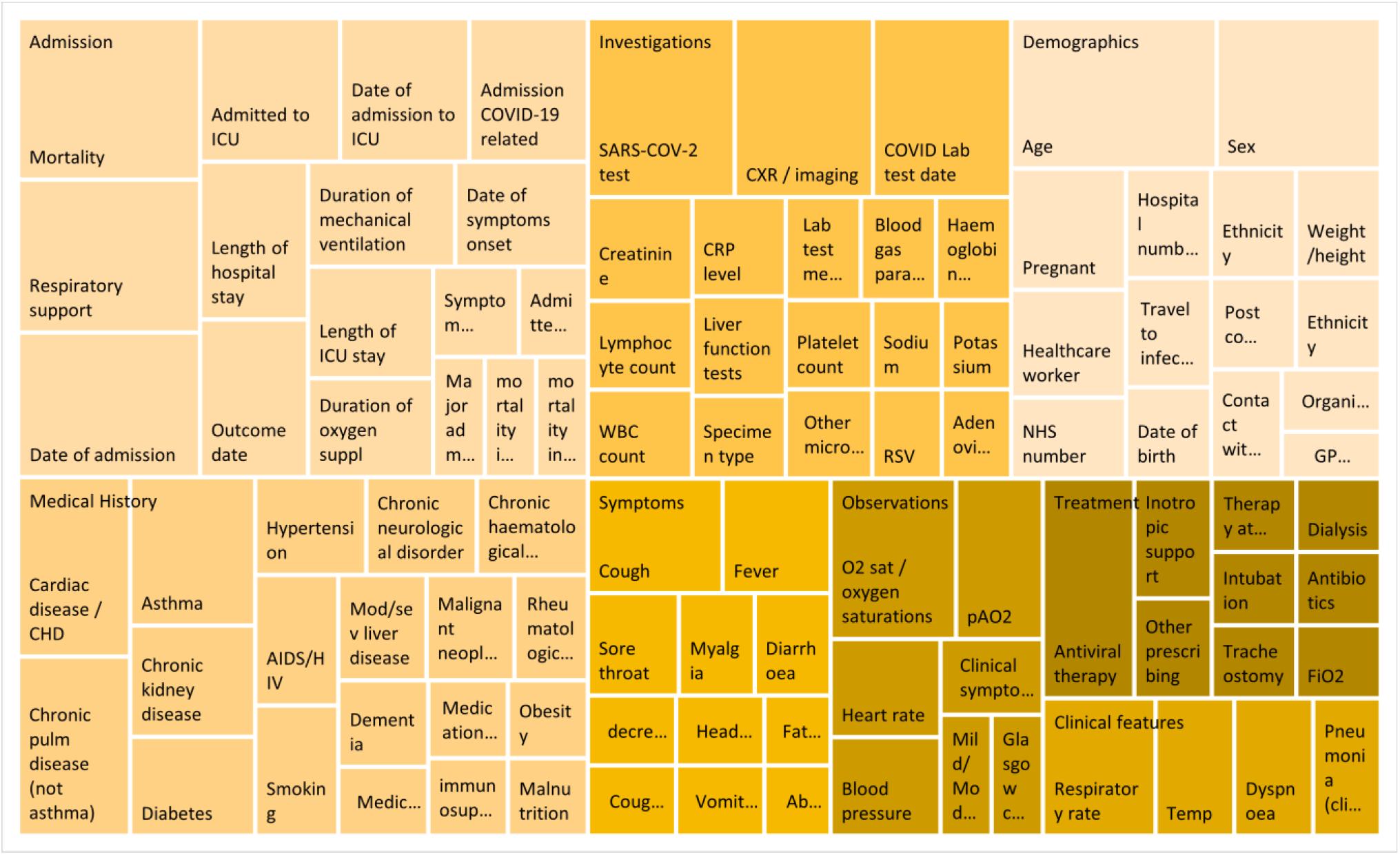
Most frequent 100 COVID-19 study data elements presented as Treemap chart in which the relative size of the rectangles represents relative frequency across studies

## Discussion

The findings of this study have demonstrated that it is feasible to collate specific data elements recorded across a range of studies investigating a specific clinical condition in order to identify those elements which are likely to be common among studies, regardless of the specific intervention or treatment which is being evaluated. In the case of COVID-19, these data may be of value for those establishing new studies, since a common set of study data elements can be rapidly developed and only highly study-specific additional data elements require addition. Furthermore, if such information were available for all recorded studies across a given clinical area, for example all of the 2000+ COVID-19 studies now in progress, an evolving set of core data elements would continually be available to both inform future study design but also allow researchers to rapidly identify other studies which may be collecting specific data of potemtial use to address a particular clinical question, and reduce the need for duplication of multiple studies collecting similar information.

The approach and data presented here represents an initial proof of concept, and ideally could be expanded by each specific data element being associated with reference to well-defined data dictionaries and associated clinical coding systems, such as SNOMED CT, in order to further improve reproducibility interoperability of data. For the purposes of the current study, data element mapping has been performed but with no attempt to evaluate the underlying definitions or vocabularies used to assign entry or inclusion for a particular data element. However, if in a particular disease area, there was the facility to develop up-to-date data element maps with links to appropriate definitions and terminologies and ontologies, this could significantly contribute to improve standardisation of data management within a specialty. In addition, in the present study no attempt was made to evaluate the underlying data model or format, but inclusion of such data if it were widely available may further improve subsequent data interoperability by providing both core data elements and underlying common data models and structures for re-use.

The main limitations of the study are that the trials and studies included represent only a small proportion of the total registered COVID-19 studies. Nevertheless, several major large UK studies are included representing a range of COVID-19 trials, and the intention of the current work was to demonstrate the feasibility of the approach and to rapidly identify the major common data elements among studies investigating COVID-19. In addition, these present data have been derived from interpretation of study protocols and eCRF documentation where formal lists of data elements were not provided, therefore it is likely that some elements have been subjectively categorised. Ideally, strict definitions are provided for each data element with clear links to open data dictionaries, but this was rarely the case. Furthermore, the vast majority of the studies included did not provide easily accessible lists of data elements along with their definitions and terminologies, meaning that some subjective interpretation was often required in terms of the specific element usage. The majority of the studies were also associated with their own eCRF, which illustrates the issue that if many studies are underway concurrently in a similar disease area and are requiring a subset of common data elements it is highly inefficient to have multiple individual eCRF instances, most of which would usually require manual data entry of some kind. A far more efficient approach would be to collect the common data elements through existing routine hospital information systems, but in general this approach is only used by a small minority of clinical studies in general.^27^

It should be emphasised that these findings are not intended to represent either a suggested common dataset or outcomes set, but merely aim to present data regarding which data elements are most common across a selection of COVID-19 studies. Indeed, for particular purposes, expert groups establishing common outcome measures in relation to COVID-19 and other diseases are underway (and indeed some of the common outcome sets included here).^28^ Whilst the findings presented are however, intended to be used as a general guide for researchers and those developing future COVID-19 studies, details for specific studies should be obtained directly from the source reference. Finally, the data presented here represent an initial group of studies investigating COVID-19, as a proof of principal in relation to the opportunity of a novel disease pandemic, requiring a rapid number of clinical studies to be set up investigating common condition. Despite the current emphasis on COVID-19, large numbers of studies are underway across all areas of medicine, and the principles illustrated would provide even greater value for the large number of studies investigating common conditions such as cardiovascular disease and cancer. The establishment of systems in which all clinical studies investigating a particular condition or within a specific domain would be required to define their data elements, and record such elements using well described standards with the aggregated data available in an open platform, would provide an invaluable resource for research planning and would significantly contribute to improved research data management and the facilitation of true FAIR data.^2^

## Data Availability

Data can be obtained via HDRUK directly. All data element details are available publicly through the referenced study websites and contacts

## Acknowledgements

This work was supported by Health Data Research UK, an initiative funded by UK Research and Innovation, Department of Health and Social Care (England) and the devolved administrations, and leading medical research charities

